# The Paradigm Shift: Reclassification of SARS-CoV-2 and Longitudinal Immune Responses in Japanese Healthcare Workers

**DOI:** 10.1101/2024.01.18.24301513

**Authors:** Tokuhiro Chano, Hiroko Kita, Tomoko Yamashita, Hirokazu Fujimura, Toshiyuki Ikemoto

## Abstract

Under the aegis of the National Infectious Diseases Act, the virus transitioned from a Category 2 menace to Category 5, analogous to seasonal influenza. The current study has investigated the protracted immune responses of healthcare workers (HCWs) towards SARS-CoV-2 in Japan. One year subsequent to the systematically implemented SARS-CoV-2 vaccination campaign among HCWs, humoral and cellular immune responses were sustained at levels as high as or higher than those immediately following the 3rd booster vaccination. Persisting immunity has highlighted the resilience and lasting memory exhibited in HCW defense against the virus.

---

In Japan, as of May 8, 2023, pursuant to the Japanese National Infectious Diseases Act, SARS-CoV-2 underwent a reclassification from Category 2, akin to SARS or highly pathogenic influenza, with profound implications for human well-being and public health, to Category 5, aligning it with seasonal influenza. Within the ambit of Category 5, no administrative measures or imperatives for vaccination or prevention of SARS-CoV-2 infection exist, leaving the onus on each individual to safeguard their own health.^1^ To arrive at this juncture, the coronavirus disease 2019 (COVID-19) must, by implication, be equated to a common cold or seasonal flu.

In the course of our surveys, the initial one transpired just prior to the commencement of the SARS-CoV-2 vaccination initiative for healthcare workers (HCWs). Consequently, the serological antibody levels of the participants remained unaffected by the vaccine.^2^ The subsequent survey occurred seven to eight months subsequent to the inaugural vaccination campaign, i.e., antecedent to the administration of the 3rd dose of the vaccine and amidst the progression of the sixth and seventh waves (Omicron strain-predominant epidemics) in Japan. This survey provided insights into humoral and cellular immune sustenance following repeated vaccinations in HCWs, up to 4 months after the 3rd vaccination.^3^

The current study, conducted at the end of October 2023, transpired 20 months subsequent to the 3rd booster dose and approximately 1 year following the SARS-CoV-2 vaccination initiative, systematically implemented in HCWs until October in the autumn of 2022. We have delineated the assessment of humoral (IgG to the Receptor-Binding Domain (RBD) of the spike protein, SARS-CoV-2 IgG II Quant, Abbott ARCHITECT®; Spike-IgG) and cellular (T-SPOT® Discovery SARS-CoV-2 kit, #DISCOVERY.432, Oxford Immunotec; T-SPOT value) immunities among 48 HCWs under continual observation. This HCW cohort exhibited a 60.4% female predominance (19/29) and was characterized by an average age of 46.6 ± 11.3 years and a body mass index (BMI) of 22.0 ± 2.5 kg/m2. In this investigation, the HCW population received vaccination on an average of 4.2 ± 0.9 occasions, with the blood collection timeframe being 12.1 ± 5.3 months from the administration of the last dose of the mRNA vaccine, which was a bivalent formulation (Omicron BA.4 or BA.5 + Wuhan-Hu-1 strain). Twenty-six cases (54.2%) had a prior diagnosis of COVID-19, and the analyzed timing from the diagnosis was 12.2 ± 5.8 months (appendix 1-2).

Abbott Spike-IgG values recorded immediately before, after, 4 months subsequent to the administration of the 3rd booster dose, and approximately 20 months, corresponding to about 1 year after the 4th vaccination, the current investigation, were; 921.7 ± 845.4, 22,903.1 ± 17,820.8, 11,494.4 ± 14,570.4, and 26,457.1 ± 28,270.3 (AU/mL), respectively. Even a year post the systematic vaccination campaign, many HCWs were observed to maintain humoral immunity at levels comparable to those immediately following the 3rd booster vaccination (figure A). The extent to which elevated spike-IgG values contribute to neutralizing antibodies against concurrently circulating omicron subvariants and their efficacy in preventing infectivity remains questionable. However, the HCWs population likely attained a state of certain herd immunity. On the other hand, T-SPOT values for SARS-CoV-2 peptides at the respective time points were; 85.0 ± 84.2, 219.4 ± 230.4, 111.1 ± 133.6, and 310.0 ± 337.2 (SFU/10^6^ cells), respectively. Cellular immunity in the current HCW cohort was seemingly sustained at levels as high as or higher than those immediately following the 3rd booster vaccination. Additional T-SPOT values for nucleocapsid and membrane proteins appeared to have occurred through natural infection of SARS-CoV-2 both pre and post vaccinations (figure B). Although 54.2% (26/48) of the cases had received a diagnosis of SARS-CoV-2 infection in alignment with questionnaire results, 81.3% (39/48) of the cases exhibited reactivity to nucleocapsid and/or membrane proteins in the T-SPOT evaluation. The duration until the present study for 10.4% (5/48) of those with common cold symptoms, possibly indicative of COVID-19 without a formal diagnosis, was 2.6 ± 1.5 months (appendix 1-2), suggesting a continuous occurrence of repeated coronavirus exposure in the post-vaccinated HCW population, possibly due to SARS-CoV-2 subclinical infection and/or common cold-like coronaviruses. Anti-spike antibody titer evaluation and the natural infection rate using positivity of anti-nucleocapsid antibody have been analyzed in each individual and/or Japanese population, and just under half are reported to have spontaneously contracted SARS-CoV-2.^4^ The evaluation of immunocompetence, encompassing the maintenance of cellular immunity, remains incomplete. However, a recurring exposure to coronaviruses in the HCW population is postulated, forming a state of herd immunity with additional cellular immune memories responsive to nucleocapsid and membrane proteins of SARS-CoV-2.

Over the 20-month follow-up of 48 HCWs, none of the documented COVID-19 cases (26/48) progressed to severe illness, and no instances of long-COVID were reported. This includes individuals with common cold symptoms (5/48), with only 14.6% (7/48) requiring more than 3 days off work, and the majority experiencing mild illness. Only two cases out of 48 HCWs with T-SPOT <20 and Spike-IgG <6,000, whose immune status could not be affirmed as enough, involved one with renal failure and peritoneal dialysis and another with asthma (appendix 1-2). In light of the aforementioned findings, it is posited that SARS-CoV-2 infection itself, among the Japanese HCW population, has evolved into a condition akin to that of the common cold or seasonal influenza. However, it is crucial to note that the actual Omicron variant infection carries a higher mortality rate than seasonal influenza or Respiratory Syncytial Virus infections.^5^ Furthermore, a recent subvariant of Omicron, BA.2.86, appears to possess a heightened ability to infect human lung cells compared to other variants, thereby elevating the likelihood of more severe lung involvement.^6^ Thus, the repeated vaccination should be recommended in high-risk cases among HCWs.

In summary, the present survey data suggests that the categorization of SARS-CoV-2 infection as a Category 5 in Japan, where no administrative measures or requirements for vaccination or prevention are imposed, appears to be appropriate.

## Data Availability

All data produced in the present study are available upon reasonable request to the authors.

## Ethical statement

Written informed consent was obtained from all study participants. During the process of obtaining the consent, all participants were informed of the need to publish the results. All participants’ personal information was anonymized, and no participant is identifiable in the report. The Research Review Board of the Shiga University of Medical Science checked all the processes and contents of this series of studies in accordance with relevant laws and regulations, and approved (No. R2021-039).

## Funding and Competing interests

This work was funded from the budget of Department of Clinical Laboratory Medicine, Shiga University of Medical Science. The authors declare that no competing interests exist.

**Figure:**
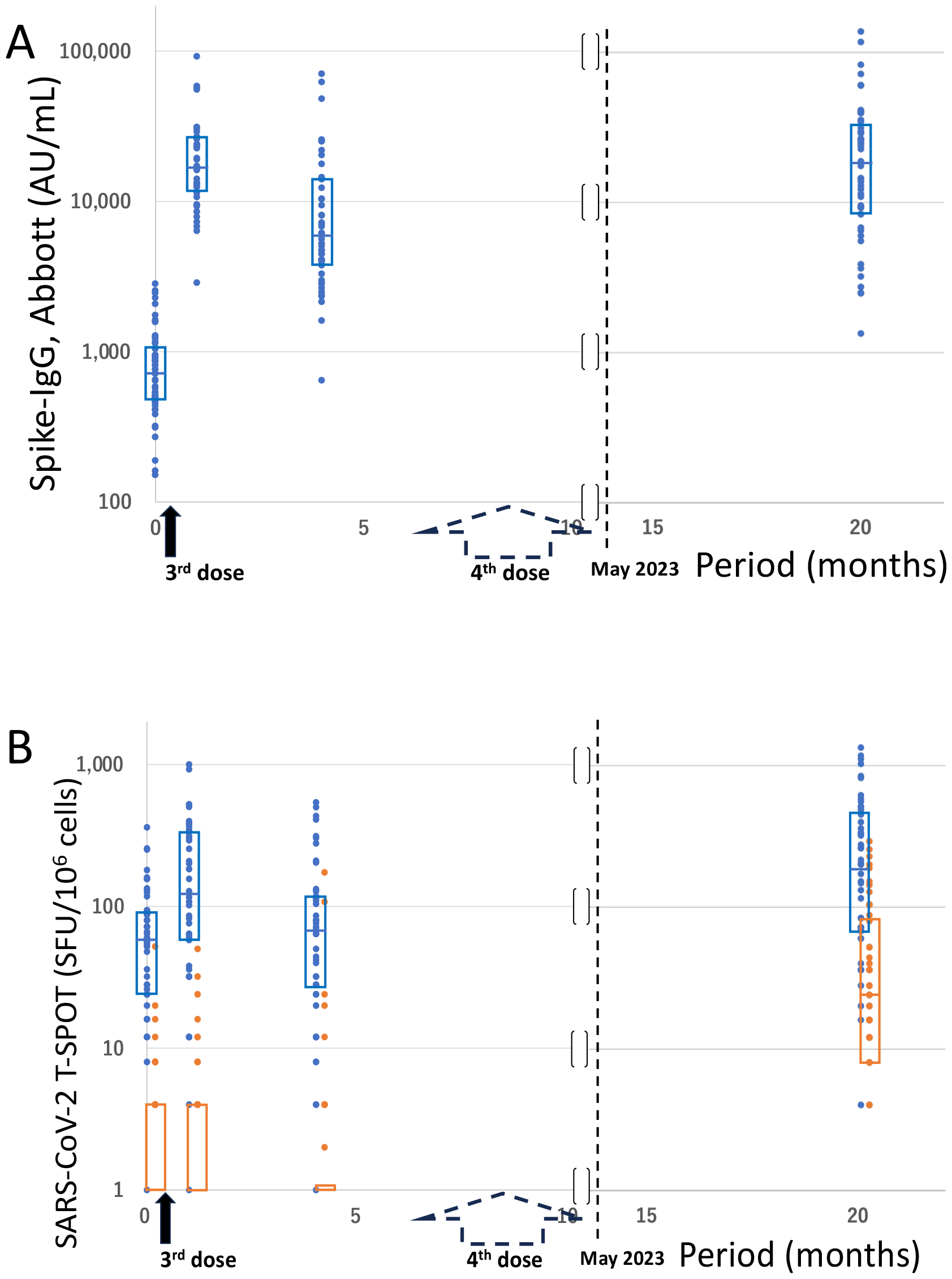
Time-elapsed values of Spike-IgG and those of T-SPOT for SARS-CoV-2. Timings of the 3^rd^ and 4^th^ vaccination dose are indicated on the X-axis (months). The black and dotted arrows indicate the 3^rd^ and 4^th^ vaccines, respectively. On the Y-axis, the Spike-IgG values (SARS-CoV-2 IgG II Quant, Abbott; AU/mL) and T-SPOT values for SARS-CoV-2 peptides (Oxford Immunotec; SFU/10^6^ cells) are indicated in **A** and **B**, respectively. The 48 healthcare workers (blue dots) were periodically followed up immediately before, after, 4 months subsequent to the administration of the 3rd booster dose, and for up to 20 months, corresponding to about 1 year after the 4th vaccination. In the T-SPOT evaluation (**B**), the values (orange dots) for non-vaccine-derived nucleocapsid and/or membrane proteins were also analyzed. Each dots represent the measured values of individual cases, and boxes in each dot section indicate the inter quartile ranges. *Note*: SARS-CoV-2, severe acute respiratory syndrome coronavirus 2; IgG, immunoglobulin G.

**Appendix 1**. Individual questionnaire for assessing the health conditions of participants, before and after SARS-CoV-2 vaccination. (xlsx)

*Note*. SARS-CoV-2, severe acute respiratory syndrome coronavirus 2

**Appendix 2**. Individual data regarding T-SPOT values representing cellular immunity against SARS-CoV-2 and the antibodies and health conditions of 48 healthcare workers, followed up periodically for up to 20 months of the 3rd dose, corresponding to about 1 year after the 4th vaccination in Shiga Prefecture. (xlsx)

*Note*. SARS-CoV-2, severe acute respiratory syndrome coronavirus 2

